# Automated outbreak detection systems in the EU: Requirements and challenges for its implementation, 2023/2024

**DOI:** 10.64898/2026.02.20.26346630

**Authors:** Paulina vom Felde genannt Imbusch, Ann Christin Vietor, Inessa Markus, Michaela Diercke, Alexander Ullrich

## Abstract

Automated outbreak detection can enhance infectious disease surveillance by enabling early identification of outbreaks and supporting timely public health measures. However, information on its current use by national public health institutes (NPHI) remains limited. This paper provides an updated and extended overview of automated outbreak detection usage in the European Union (EU) and United Kingdom (UK). Key findings were gathered through the Joint Action United4Surveillance via an online survey of 21 countries, an in-presence workshop, and online meetings with NPHI, focusing on three objectives: assessing current demand for automated outbreak detection, examining the availability of necessary prerequisites within existing surveillance systems, and identifying challenges and requirements for implementation. Findings indicate that seven countries currently have automated outbreak detection systems (AODS) in place. While many countries have sufficient surveillance data and a clear demand for automated outbreak detection, adoption is often limited by constrained funding and lack of IT resources. While the specific methods in existing AODS differ, overall demands and outputs are similar, suggesting a single tool could serve multiple countries. Capacity building as part of EU-funded Joint Actions can bridge these gaps by developing sustainable tools and fostering cross-country collaboration.

## Introduction

Timely detection of infectious disease outbreaks, enabling targeted investigations and control measures, is a core objective of public health surveillance [1, 2]. Outbreak detection relies on identifying case numbers that exceed the expected threshold, typically recognised through accumulated case reports of notifiable diseases or outbreak notifications from reporting entities. With the growing availability of electronic case reports, automated outbreak detection has gained significant importance, offering a systematic approach to identify outbreaks [1, 3, 4].

Automated outbreak detection refers to the process of identifying unusual increases in case numbers by comparing the current epidemiological situation with a baseline, usually established from historical or geographical data. Statistical methods estimate the expected number of cases and compute a threshold. When observed values deviate significantly from these expectations, surpassing the threshold, signals are generated and often presented in tables, visualisations, or descriptive reports. These signals necessitate epidemiological investigations to determine whether they represent potential outbreaks or false positives [5].

Various methods for outbreak detection exist and have evolved over time [6–8]. Among the most widely used is the method by Farrington and adaptations by Noufaily, which employ generalised linear models to predict the expected number of cases, accounting for seasonality, time trend and overdispersion [9, 10]. Building on these methods, we can distinguish a hierarchy of components: statistical methods underlie tools that operationalise them and some of these tools allow the automated analysis of complete surveillance datasets. Together with the necessary infrastructure, standard operating procedures (SOPs), and trained personnel, these tools constitute comprehensive automated outbreak detection systems (AODS).

Despite methodological advances, information on AODS implementation in the European Union (EU) is limited. The most recent overview from 2010 reported six institutes in five countries (Denmark, Germany, the Netherlands, Sweden and the United Kingdom [UK]) using automated detection for routine surveillance. Based on the existing AODS and practical experiences, recommendations including the regular evaluation of AODS and aiming for a user-friendly approach directed at epidemiologists were drawn [11, 12]. Additionally, a 2017 workshop involving stakeholders from 17 institutions across the EU emphasised exchange of data and experiences across countries, the need for visual outputs for users, and assessment and comparison of detection methods [13].

One aim of the EU4Health Joint Action UNITED4Surveillance (U4S) was to identify the requirements of EU countries for effective outbreak detection [14]. Understanding these needs and current practices is a necessary first step before developing and implementing new AODS. While the project also involved developing and piloting an open-source R Shiny app for automated outbreak detection by multiple NPHI, this paper provides an updated and extended overview of automated outbreak detection usage in EU countries and the UK. The analysis encompassed (1) assessing current demand for automated outbreak detection, (2) examining availability of necessary prerequisites of surveillance systems, and (3) identifying challenges and requirements for running AODS.

## Methods

A mixed-method approach was employed, combining a survey, a workshop and online meetings as part of the U4S project. The UK and France did not participate in the survey but were included as additional case studies because the 2010 overview and the workshop, described in the introduction, identified these countries as operating relevant AODS [11, 13]. In the 2010 overview, UK systems were reported separately for England, Wales and Northern Ireland. Therefore, our analysis distinguishes these jurisdictions where relevant.

### Questionnaire development

A questionnaire was drafted with the goal to identify gaps and needs from the perspective of epidemiology and data science respectively. Respondents were asked to select one national surveillance system either in current use or consider most suitable for automated outbreak detection. The questionnaire had two sections: (1) description of public health surveillance systems including types, legal frameworks, disease groups, data sources, aggregation level, reporting frequency, indicators, geographical resolution, and historical data availability; and (2) automated outbreak detection capacities, covering resource availability, signal follow-up, and details of existing methods, resources, and outputs.

The draft was reviewed by six project partners and a contact point at the European Centre for Disease Prevention and Control (ECDC) and revised accordingly. The final version (Supplementary Material S1) contained 33 questions of which 22 questions were mandatory, primarily single or multiple-choice rankings and free text fields.

### Survey administration

The survey was created using the EUSurvey platform and distributed on 17 April 2023 by the U4S coordinator to designated contact points of the 25 countries participating in the project. As multiple experts (epidemiologists, data scientists) were needed to complete it, contact points were encouraged to consult national experts.

### Workshop

An onsite workshop was held in May 2023 at the RKI in Berlin, Germany, with 30 public health experts, epidemiologists, and data scientists from NPHI of 16 EU countries as part of the U4S project. The workshop aimed to discuss survey findings, identify relevant use cases, and plan the pilot phase. Participating countries presented existing AODS and surveillance systems overviews. Participants were divided into two breakout groups. Data scientists worked on defining outbreak detection tools achievable within the next 12 months, including requirements, vision, data input/output, and handling (operation, users, interfaces, reporting). Epidemiologists as the future tool users worked on outlining the ideal tool, specifying intended functionality, data sources, pathogens, use cases, and operational aspects.

### One-on-one meetings and calls

In January 2024, one-on-one meetings were conducted with the ten piloting countries. These sessions focused on gathering detailed requirements for the tool, including country-specific administrative divisions and individual functional needs. Additional informal online meetings conducted between 2023 and 2024 further supported the collection and clarification of tool requirements.

### Data analysis and verification

Data are presented in a descriptive manner. Survey answers were processed and analysed using R (version 4.4.0) [15]. Survey results were first presented during the workshop in May 2023 allowing participants to review and validate data. During a project webinar in June 2023 the survey and workshop results were shared with the entire project consortium, giving all survey respondents the opportunity to verify their data.

## Results

The analysis included 26 countries, comprising 25 EU member states (MS) and the UK. Among the 25 U4S consortium members invited to participate in the survey, 21 completed it, yielding a response rate of 84%. The largest group of countries (n=16; 62%) contributed data through the survey and participation in the workshop. Five countries (19%) participated in the survey only, data for two countries (8%) were obtained solely from literature, and for three countries (12%) no data were available (Figure 1). The majority of survey respondents are associated with NPHI (n=19; 90%) and the remaining with Ministries of Health (n=2; 10%).

**Figure 1.**
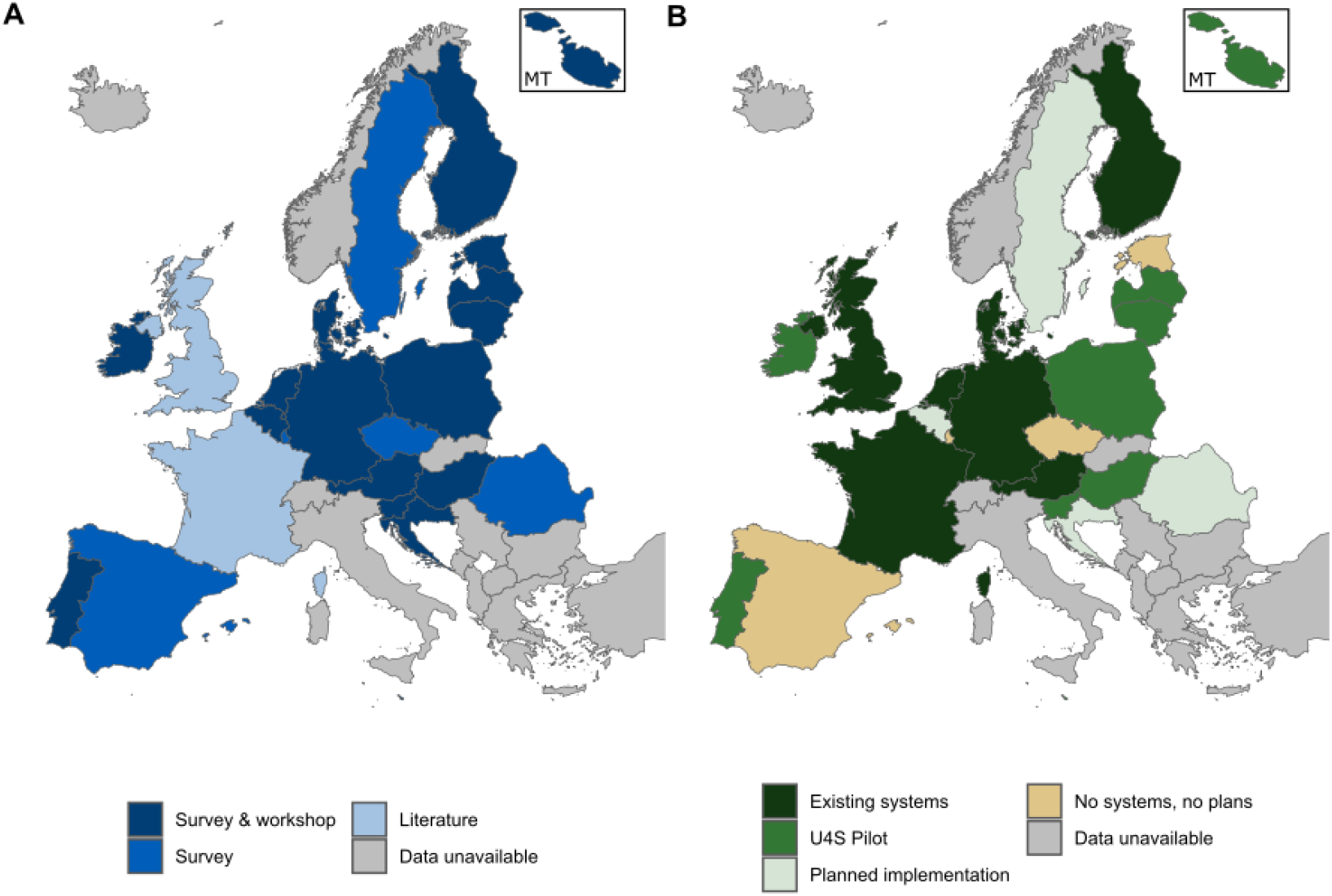
Map showing the current status of AODS in the EU + UK and the corresponding source of information, 2024. **A:** Source of information used to assess the status of AODS across the EU + UK. All countries that participated in the workshop also completed the survey. The upper right inset shows Malta (MT). **B:** Countries are categorised to their current level of implementation of AODS. Where countries meet criteria for multiple categories (e.g. Finland, Denmark and the Netherlands, which have existing AODS and are also U4S pilot partners), the higher level of implementation is displayed. U4S Pilot refers to those countries who used the automated outbreak detection tool, developed within U4S, in a pilot phase from April-December 2024. The upper right inset shows Malta (MT).

### Current usage of AODS in EU countries

Based on the collected data from EU countries and the UK, seven countries have existing AODS in place. Ten countries participated in the U4S pilot project, while four countries are planning to implement such AODS. Another four countries have neither AODS nor plans for implementation, and for three countries no data is available (Figure 1).

### Identified need for outbreak detection

Our survey examined how countries respond to signals generated from surveillance data, whether detected manually or automatically. While a diverse level of implementation across the EU is observed, all 21 countries have stated in the survey that they would act on a signal that is found in their surveillance data, whether from manual inspection or automated analyses. Typically, this response involves verification of the signal, followed by the initialisation of an investigation, a risk assessment, further communication and introduction of measures if needed. In most cases (n=14) the respective NPHI receives feedback on actions triggered by a signal, either via outbreak reports (n=6), direct communication with the investigation authorities (n=3), or by observing the performed public health action (n=2). In one case, investigations were conducted directly by the NPHI (n=1). In many countries (n=11) feedback on actions is collected systematically and often also used for evaluation (n=8). In addition, piloting countries grew from initially seven to ten, reflecting the high interest in the tool.

### Identified challenges for implementing AODS

When asked about main challenges in implementing AODS, lack of sustainable funding and resources were indicated as the primary obstacle (n=15). This was followed by data quality (n=9) and legal mandate issues (n=5). In one country, data quality specifically referred to limited data availability and reporting delays. Two countries reported no barriers, while another two countries reported ‘other’ issues. Of these, one respondent (Sweden) noted that an automated detection system had previously been implemented in the surveillance system but was not yet re-implemented following a system upgrade in 2022. The other country cited concerns about potential functional incompatibilities between systems, referring to technical issues that can cause delays or inconsistencies when data are synchronised across platforms before integration into the national eHealth system.

Regarding funding, respondents reported particularly limited funding of research capacity, specifically staff knowledge and time for research activities (n=12), personnel (n=8), overall funding (n=7) and IT resources such as hardware and software (n=6). Similarly, no funding was available for IT competences, including research software engineering for implementation (n=6), IT support for maintenance and automatization (n=6), and data science expertise for development and extension (n=8) (see Figure 2).

**Figure 2.**
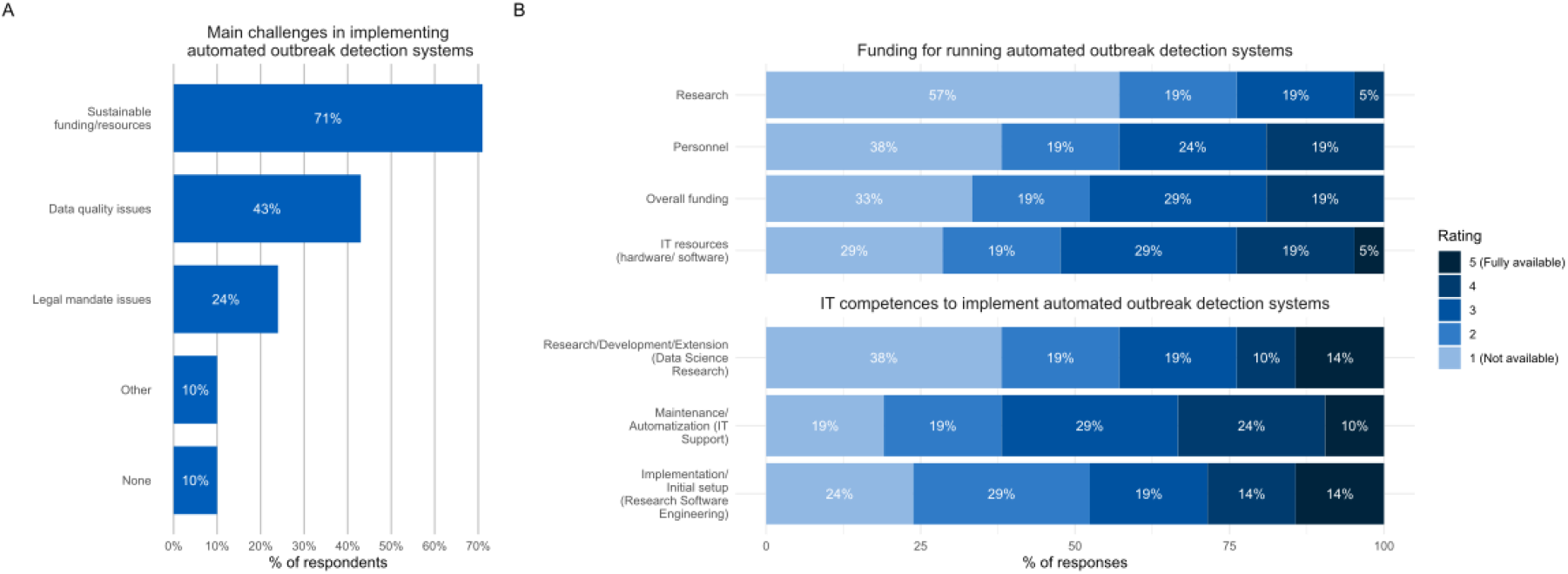
Reported challenges and resources for implementing AODS in the EU, 2023. **A:** Main challenges reported by countries in implementing AODS. Multiple answers per respondent possible. **B:** Aggregated ratings (1-5) of funding availability and IT competences to run and support implementation. Higher scores indicate greater perceived availability.

### Surveillance systems prerequisites

The performance of AODS primarily relies on the information available in the surveillance system, including historical data used to establish baselines, detect trends, and adjust for seasonality [5]. Almost all respondents stated that data is reported case-based (n=20) and on a daily basis (n=19) to the national level. Among these, 12 stated that additional personal identifiable data (e.g. address, date of birth, personal identity number) is available. Further, all countries (n=21) have at least 3 years of historic data available, 8 countries have historic data for 20 years or more.

All countries (n=21) receive information on age, region, pathogen and sex of infectious disease cases through their surveillance systems at national level. These are the most common stratification dimensions in AODS and allow for extensive investigation. Most countries also have additional information on outcome like death (n=19) and hospitalisation (n=17) and date of symptoms onset (n=17) or specific symptoms (n=14), which would allow for further assessment of signals regarding the severity or even outbreak detection on different indicators (e.g. number of hospitalisations). In many countries information on vaccination status (n=15), risk factors (n=11) or exposures (n=15) exist, which would enable further disease-specific analyses.

### Identified use cases for AODS

Identifying potential use cases for automated outbreak detection is essential for designing an effective and widely used AODS. This includes specifying data input formats, defining user-configurable input options such as filters and parameters and ensuring that analysis and outputs are actionable and relevant for various disease surveillance systems [16].

The survey collected information on disease groups targeted by the surveillance systems, revealing potential use cases for AODS and groups that could benefit most from these systems. Food- and waterborne diseases (n=19), air-borne diseases (n=18) and vector-borne disease and zoonotic disease (n=17) were the most common disease groups. At the workshop, gastroenteritis and respiratory diseases emerged as the main use cases, with stronger interest in gastroenteritis. Accordingly, pilot use cases focus mainly on gastrointestinal pathogens, with one other country selecting Group A streptococci (GAS).

### Identified requirements for AODS

The identified requirements for AODS drawn from one-on-one meetings calls with the piloting countries, show significant overlap despite different organisational and geographical structures. Key needs include flexible output formats, adaptability for pandemic periods and various filtering and stratification options. The systems should also provide information on cases related to already known outbreaks, enable extraction of detailed case line lists, support different temporal aggregations like daily, weekly and monthly, and offer multiple language options. The workshop discussion highlighted that signals are managed by different experts across countries with varying levels of IT expertise, underscoring the need for user-friendly, accessible, and adaptable tools. These requirements highlight the need for a flexible system that can accommodate diverse user needs effectively.

### Current AODS implemented by countries

Table 1 presents an overview of current AODS implemented in seven countries. The results are presented in two paragraphs highlighting the differences in type and depth of information between countries that participated in the U4S project and France and the UK, for which only literature-based data were available.

**Table 1.** Overview of AODS in six EU countries and the UK.

| Country | Pathogens;<br>Surveillance data | Method(s) | Implementation;<br>Automation | Reporting;<br>Frequency | Remarks |
| --- | --- | --- | --- | --- | --- |
| **Austria** | Campylobacter & Salmonella;<br>Case data from the national reference laboratory | Farrington Flexible, CUSUM | R, surveillance package; fully automated | Weekly report | COVID-era hiring added data-scientist capacity; plan to use data from national electronic reporting system and to extend to all notifiable diseases |
| **Denmark** | ~20 assorted notifiable diseases;<br>Campylobacter, COVID-19;<br>Lab data from national microbiology DB | Threshold rules for ~20 diseases;<br>Farrington Flexible, in-house method for COVID-19 prevalence levels | SQL + R + SAS, surveillance package; automation varies | Daily auto email alerts (~20 diseases); weekly ad-hoc R reports or web-figure | No single framework; evaluation pending; initiative to unify |
| **Finland** | Influenza;<br>Cases from Register of Primary Health Care visits | Moving Epidemic Method (MEM) | R, mem package; fully automated | Daily website updates on case numbers, thresholds & epidemic levels | MEM since 2015; focused on influenza; plan to monitor other infectious diseases with data from National Infectious Diseases Register |
| **Germany** | ~30 diseases; National case reports from German surveillance DB | Farrington Flexible | R pipeline, surveillance package, fully automated | Comprehensive weekly report (maps, charts, tables) | Extensive coverage of notifiable pathogens |
| **Netherlands** | ~40 diseases;<br>Case-based data in RIVM DB | Hidden Markov model + spline seasonality | R-Shiny dashboard ('survboard'); AdHoc computation during usage of dashboard | Interactive dashboard; Frequency dependent on usage | Real-time geographic, temporal & demographic views; Plan to use WGS data for outbreak detection with new Shiny app |
| **France (SurSaUD<sup>®</sup>) – Syndromic** | Influenza + 11 other syndromes;<br>Emergency dept & SOS Médecins visits + mortality (~700 EDs, 60 GP units), >90% ED coverage | MASS: Ensemble of GLM, robust GLM (weighted) & Hidden Markov model | Daily R scripts at Santé Publique France; alarms fully automated, human validation of alert levels | Daily national dashboard & weekly reports | performance benchmarked comparing ensemble vs. individual methods and application to different data sources |
| **UK (England) – Syndromic** | Comprehensive coverage of syndromes; NHS 111 calls, GP consultations, ED attendances | RAMMIE (rising-activity multi-level mixed-effects indicator emphasis) | automated Stata script; daily | Daily internal alerts and public weekly bulletins | RAMMIE demonstrated high sensitivity ($\approx 92\%$) & specificity ($\approx 99\%$); adjusts for weekday/holiday effects |
| **UK (England & Wales) – Lab isolates** | >3300 distinct organisms; HPA surveillance data | Improved Farrington (quasi-Poisson GLM) | R scripts (surveillance package); fully automated | Weekly reports emailed to national and regional epidemiologists | One of Europe's oldest automated systems; continuously refined |

Austria, Denmark, Finland, Germany, and the Netherlands all use routinely collected notifiable infectious disease surveillance data and make use of R-based workflows including the surveillance R package [17]. They differ, however, in the number and type of pathogens monitored, the specific outbreak detection method applied, the degree of automation and the type of reporting. The number of pathogens monitored varies from a single pathogen (Influenza; Finland), two pathogens (*Campylobacter*/ *Salmonella*; Austria) to a comprehensive coverage of 30-40 notifiable diseases (Germany, Netherlands). In terms of outbreak detection methods, Austria, Denmark and Germany use the FarringtonFlexible [10]. Finland adopts the Moving Epidemic Method (MEM) for Influenza and the Netherlands applies a custom hidden Markov model [18]. Despite this methodological heterogeneity, all systems generate near-real-time outputs: daily updated dashboards (Finland), fully automated weekly reports (Austria, Germany), auto-email alerts and weekly figures (Denmark) as well as the usage of an interactive Shiny app on the latest data (Netherlands).

In Denmark, several parallel scripts and legacy SAS-based solutions remain in use, lacking a harmonised, institute-wide framework for automated outbreak detection. Austria and Finland are aiming to extend their systems to cover a broader range of notifiable diseases and to use the national infectious disease data, either as a replacement for or in addition to the data sources currently in use.

AODS in France and the UK are applied on diverse national data streams and use established R-based analytic frameworks. In both countries, there is a comprehensive coverage of syndromes (France & UK) or pathogens (UK). In France, the syndromic surveillance system SurSaUD utilizes emergency department attendances, SOS Médecins consultations and mortality data and applies an ensemble of methods, namely GLM, robust GLM and a hidden Markov model [19]. In the UK, the syndromic surveillance system incorporates NHS 111 calls, GP and ED attendances and uses the RAMMIE method [20], while outbreak detection for laboratory surveillance uses FarringtonFlexible [10]. In both countries automated R scripts run daily or weekly, producing internal alerts, dashboards and bulletins. The systems in France and the UK have been established for many years, reflect a high level of methodological maturity and offer guidance for other countries through systematic evaluations of the used methods.

## Discussion

This paper provides an updated overview of the current landscape of AODS in the EU and the UK. Our findings reveal a heterogenous state of implementation, with only seven countries included in the analysis having established AODS. However, our results also highlight important opportunities for broader adoption, given the availability of rich surveillance data across many countries and meeting key prerequisites for automating processes.

Compared to the 2010 overview, which outlined AODS in Denmark, England, Germany, the Netherlands and Sweden, our findings show that the overall landscape has changed little [11]. Most of these countries continue to operate AODS, except Sweden, which discontinued its systems following the implementation of a new surveillance system. Austria and Finland are the only countries that have introduced new systems: Austria’s enhanced capacities during the COVID-19 pandemic may reflect increased prioritisation of AODS, enabled by the additional resources made available during the pandemic. In contrast, Finland’s adoption using the MEM method focuses more on retrospective trend analysis than real-time signal detection. The findings also suggest that high-resource countries are more likely to establish outbreak detection systems, but maintaining them can become challenging when substantial changes of surveillance systems occur.

Limited funding and IT capacity remain the main barriers to wider implementation. Deficits in software engineering, IT support, and data science restrict the ability of NPHIs to develop and sustain systems. These constraints help explain why, despite clear interest, only a minority of countries currently operate AODS. At the same time, surveillance environments are evolving. An increasing number of countries are routinely generating whole genome sequencing (WGS) data as part of surveillance activities, which introduces new requirements for outbreak detection tools, particularly with regard to data input formats, interoperability, and the ability to integrate sequencing information alongside epidemiological data.

Our survey and pilot project further indicate growing momentum toward automation. Ten countries actively piloted the developed tool, demonstrating both the technical feasibility of implementation and the willingness of NPHI to integrate methods into routine practices. In parallel, all survey respondents confirmed that they would act on signals detected in their surveillance data. This finding indicates that the principle of responding to signals is firmly embedded in national public health practice. Structured procedures for response are in place in most countries, yet the way feedback is collected and integrated varies considerably. Reliance on manual verification may cause delays and inconsistencies, while greater automation could enable timelier signal identification, more consistent risk assessment, and systematic feedback.

Moreover, the results show significant potential for implementing new outbreak detection system, as many countries reported having extensive historical data often exceeding eight years, which is crucial for establishing baselines and trend analyses. Identifying specific use cases, such as gastrointestinal pathogens, highlighted the practical value of AODS in routine surveillance tasks. Another potential application of new methods such as spatial clustering at the address level arises from the availability of certain personally identifiable data. The ability to incorporate such data allows for more precise mapping and detection of patterns.

Experience from pilot and established systems shows substantial alignment in outbreak detection needs across the EU. Despite differences in epidemiological contexts and organisational structures, system requirements show considerable overlap. Pilot countries highlighted needs such as flexible output formats, temporal aggregation, filtering, and multilingual support, while established systems use different statistical methods and infrastructures yet generate largely comparable outputs. The convergence of outputs despite methodological differences suggests that key outputs including signals, visualisations, maps and reports could be standardised. These shared requirements indicate that a single, flexible tool can accommodate diverse national contexts, meeting practical needs without requiring separate systems for each country. The wide-spread usage of the surveillance package is a good example of the utility of open-source tools developed by the public health community.

### Limitations

Our study has some limitations. First, the survey focused on one surveillance system per country, which might not fully capture the use of AODS across all national surveillance systems. Second, varying interpretations of automated outbreak detection could have influenced reported usage, including differences between signal detection and data visualisation. Third, reliance on self-reported data may have introduced bias or overestimation, with response depth varying by how extensively experts were consulted. Fourth, workshop participation was limited to 30 experts from 16 countries, not representing all EU contexts. Fifth, while the systems in the UK and France are still operational, the data may not reflect all subsequent updates. Despite these limitations, the study provides valuable insights into existing practices and priority areas for improvement.

### Future perspectives

Future work should focus on integrating the R Shiny app within the U4S project into local systems, expanding its use beyond the project phase, and refining methods, interfaces, and data integration. Broader adoption of such flexible, open-source solutions could accelerate cross-country collaboration and promote more harmonised outbreak detection in the EU. However, the high volume of signals produced by automated systems, often with limited specificity, can generate substantial verification workloads. This highlights the need for systematic evaluation of tools in real-world settings to assess their accuracy, usability, and added value in routine practice.

## Conclusion

AODS offer the potential to enhance routine surveillance of infectious diseases across EU countries. However, the results indicate that resource limitations and technical barriers often hinder their implementation. Projects like U4S can play a critical role in bridging these gaps by bringing together expertise and fostering collaboration among countries.

## Data Availability

All data produced in the present study are available upon reasonable request to the authors.

